# Neighborhood Deprivation and Disparities in Blood Pressure Monitoring in Patients with Intracerebral Hemorrhage

**DOI:** 10.64898/2026.05.22.26353704

**Authors:** Samuel Namian, Joel Smith, Sofia Constantinescu, Yome Tawaldemedhen, Cyprien A. Rivier, Santiago Clocchiatti-Tuozzo, Shufan Huo, Kane Wu, Rachel Forman, Victor Torres Lopez, N. Abimbola Sunmonu, Nils Petersen, Guido J. Falcone

**Author notes:** **Correspondence:** Guido J. Falcone, MD, ScD, MPH, 100 York Street, Suite 1N, Box 2080188, New Haven, CT 06511, USA.

## Abstract

**Background:** Patients in socioeconomically disadvantaged neighborhoods face barriers to care. Missing BP documentation may signal gaps in risk-factor management, a crucial component of primary and secondary prevention of intracerebral hemorrhage (ICH). We tested whether neighborhood deprivation was associated with absent electronic health record (EHR) blood pressure (BP) documentation surrounding ICH and whether absent documentation predicted subsequent uncontrolled BP.

**Methods:** We conducted a case-only study within the NIH All of Us Research Program. We included ICH survivors (ICD-10 I61.x, surviving >=1 year) with available ZIP3-based Deprivation Index. Deprivation was categorized as Privileged, Intermediate, or Deprived using cohort-based tertiles. We excluded BP measurements collected by All of Us. Outcomes were (1) absent EHR-derived BP documentation and (2) uncontrolled BP (mean systolic BP >=140 mmHg) during three windows: 1-365 days before ICH; 30-365 days and 1-5 years after ICH. Multivariable logistic regression tested associations adjusting for age, sex, and race/ethnicity.

**Results:** 1,474 ICH survivors were included (mean age 60.1, 50.4% female). Compared to privileged neighborhoods, those living in deprived neighborhoods had higher odds of absent EHR BP documentation in the year prior to ICH (OR 2.10, 95% CI 1.60-2.76; p<0.001), 30-365 days post-ICH (OR 2.82, 95% CI 2.14-3.73; p<0.001) and 1-5 years post-ICH (OR 2.81, 95% CI 2.13-3.71; p<0.001). Absence of EHR BP documentation in the year before ICH predicted uncontrolled BP 30-365 days (OR 1.97, 95% CI 1.36-2.85; p<0.001; N=888) and 1-5 years (OR 1.83, 95% CI 1.24-2.69; p=0.002; N=814) after ICH. Absence of BP documentation 30-365 days post-ICH also predicted uncontrolled BP 1-5 years post-ICH (OR 1.66, 95% CI 1.10-2.50; p=0.017; N=814).

**Conclusions:** Neighborhood deprivation is associated with persistent gaps in EHR BP documentation surrounding ICH, and absent documentation before or soon after ICH predicts subsequent uncontrolled BP. These findings highlight the need for community-level strategies that ensure equitable BP monitoring for socioeconomically disadvantaged populations.

## Introduction

Blood pressure (BP) control is essential for preventing recurrent intracerebral hemorrhage (ICH) and other vascular events after stroke.^1^ However, post-ICH BP control remains suboptimal particularly among socioeconomically disadvantaged populations, yet the mechanisms underlying these hypertension disparities are not completely understood.^2,3^ Effective hypertension management requires routine monitoring. At the individual level, sustained BP control requires longitudinal clinical follow-up. At the population level, designing targeted interventions requires adequate ascertainment of BP data across socioeconomic groups to precisely characterize disparities in BP control. While conducting a related analysis of BP disparities surrounding ICH in the All of Us Research Program, we observed that individuals from more deprived neighborhoods were substantially less likely to have electronic health record (EHR)-derived BP data available, motivating the present study. Whether gaps in BP monitoring contribute to post-ICH disparities has not been examined in a diverse national cohort. We hypothesized that neighborhood deprivation is associated with gaps in EHR BP documentation surrounding ICH and that these gaps are associated with worse subsequent BP control.

## Methods

We conducted a retrospective case-only study within the All of Us Research Program (Controlled Tier, version 8), a diverse national cohort of United States residents.^4^ We included adults over 18 years old with a first ICH identified by ICD-10 codes (I61.x) who survived at least 1 year and had a non-missing Deprivation Index. The Deprivation Index combines six American Community Survey indicators for each participant’s 3-digit ZIP code, categorized into tertiles relative to the study population (Privileged, Intermediate, Deprived).^5^ Systolic BP measurements were extracted from EHR data. Program-collected baseline measurements were excluded. We assessed two outcomes across three time windows (pre-ICH: 1-365 days before; post-ICH: 30-365 days after; long-term: 1-5 years after): (1) absent EHR-derived BP documentation and (2) uncontrolled BP, defined as mean systolic BP ≥140 mmHg among participants with BP data in a given window. Logistic regression estimated adjusted odds ratios (ORs) and 95% confidence intervals (CIs) for the association between deprivation and absent EHR documentation in each window, and separately for the association between absent EHR documentation in earlier windows and uncontrolled BP in subsequent windows, adjusting for age, sex, and race/ethnicity. The study was approved by the All of Us Institutional Review Board.

## Results

Among 1,474 ICH survivors (mean age 60.1 years; 50.4% female; 52.2% White, 23.1% Black, 17.1% Hispanic/Latino, 7.6% Other), EHR-derived BP documentation was absent in 34-62% of survivors depending on the time window and deprivation tertile (Figure 1, Panel A). In adjusted models, ICH survivors from deprived neighborhoods had approximately 2-to 3-fold higher odds of absent EHR BP documentation compared with privileged neighborhoods across all windows: pre-ICH (OR 2.10; 95% CI 1.60-2.76), post-ICH (OR 2.82; 95% CI 2.14-3.73), and long-term (OR 2.81; 95% CI 2.13-3.71; all P<0.001; Figure 1, Panel B). In the same models, self-reported race and ethnicity was not independently associated with absent EHR BP documentation (all P>0.2).

**Figure 1.**
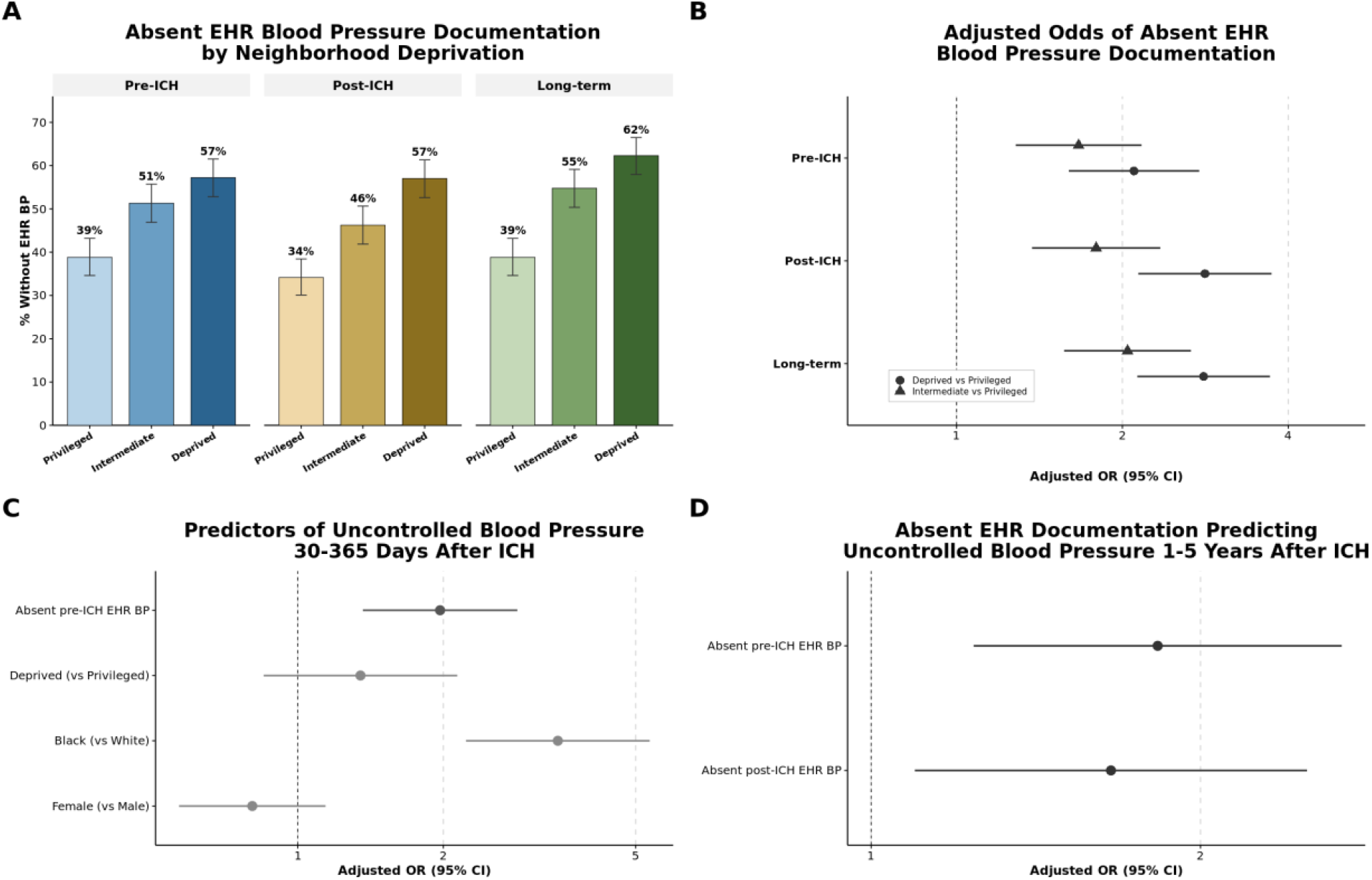
Neighborhood deprivation, blood pressure documentation gaps, and uncontrolled blood pressure surrounding intracerebral hemorrhage. *Panel A* shows the percentage of ICH survivors without electronic health record (EHR)-derived blood pressure documentation by neighborhood deprivation tertile (Privileged, Intermediate, Deprived) across three time windows: pre-ICH (1–365 days before), post-ICH (30–365 days after), and long-term (1–5 years after ICH). Error bars indicate 95% Wilson score confidence intervals. *Panel B* displays adjusted odds ratios and 95% confidence intervals for absent EHR blood pressure documentation for Intermediate and Deprived tertiles compared with Privileged in each time window. *Panel C* shows adjusted odds ratios for selected predictors of uncontrolled blood pressure (mean systolic BP ≥140 mmHg) 30–365 days after ICH (N=888), including absent pre-ICH EHR BP documentation, neighborhood deprivation, race, and sex. *Panel D* displays adjusted odds ratios for absent EHR BP documentation predicting uncontrolled blood pressure 1–5 years after ICH (N=814) from two separate models: absent pre-ICH BP documentation and absent post-ICH BP documentation. All models adjusted for age at ICH, sex, self-reported race and ethnicity, and neighborhood deprivation tertile. BP indicates blood pressure; CI, confidence interval; EHR, electronic health record; ICH, intracerebral hemorrhage; and OR, odds ratio.

Absent EHR BP documentation was associated with worse subsequent BP control. Among survivors with post-ICH BP data, those lacking pre-ICH BP documentation had nearly twice the adjusted odds of uncontrolled BP at 30-365 days (OR 1.97; 95% CI 1.36-2.85; P<0.001; n=888; Figure 1, Panel C). Among 814 survivors with long-term BP data, absent pre-ICH documentation (OR 1.83; 95% CI 1.24-2.69; P=0.002) and absent post-ICH documentation (OR 1.66; 95% CI 1.10-2.50; P=0.017) were each associated with uncontrolled BP at 1-5 years in separate adjusted models (Figure 1, Panel D).

## Discussion

In this diverse cohort of ICH survivors, neighborhood deprivation was associated with absent BP documentation in EHR before and after ICH, and absent BP documentation was associated with worse subsequent BP control. These findings suggest that gaps in BP monitoring may represent one pathway linking socioeconomic disadvantage to uncontrolled BP after ICH. These gaps also have broader implications for surveillance. If the populations most likely to have uncontrolled BP are the same populations least visible in EHR-derived data, existing estimates of post-ICH hypertension disparities may underestimate its true burden in deprived communities. After mutual adjustment, absent documentation appeared more closely related to neighborhood deprivation than to race and ethnicity, suggesting that interventions based solely on race and ethnicity may miss modifiable location-based barriers to healthcare access. Community-level strategies that improve healthcare access in disadvantaged neighborhoods may help close monitoring gaps, improve BP control, and enable more precise characterization of post-ICH hypertension disparities.^3^

Limitations include the observational design, reliance on EHR-linked BP measurements that may not capture monitoring not accessible in All of Us, and the use of 3-digit ZIP-linked deprivation which lacks granularity for precise neighborhood context.

## Non-standard Abbreviations and Acronyms

BP: Blood Pressure
CI: Confidence Interval
EHR: Electronic Health Record
ICH: Intracerebral Hemorrhage
OR: Odds Ratio

## Acknowledgments

We gratefully acknowledge All of Us participants for their contributions, without whom this research would not have been possible. We also thank the All of Us Research Program for making data available to the research community.

## Data Availability

Data used in this study are available to authorized researchers through the All of Us Research Program Researcher Workbench (https://www.researchallofus.org).

